# Understanding end-of-life multimorbidity: An analysis of multiple causes of death in Denmark

**DOI:** 10.64898/2026.07.03.26357007

**Authors:** Cosmo Strozza, Elizaveta Ukolova, Marie-Pier Bergeron-Boucher

**Affiliations:** Interdisciplinary Centre on Population Dynamics, University of Southern Denmark

## Abstract

**Background:** Mortality analysis traditionally focuses on the single underlying cause of death (UCD), which obscures the wider morbidity process at the end of life. Multiple causes of death (MCoD) data, recording all conditions on the death certificate, are increasingly used as a proxy for end-of-life multimorbidity, yet how accurately they represent it remains underinvestigated. We assessed whether recorded causes reflect end-of-life health conditions or rather the chain of events leading to death.

**Methods:** Using linked Danish registers (Population, Cause of Death, Chronic Diseases, and Cancer), we studied residents aged 50+ diagnosed with COPD, dementia, diabetes, or cancer who died in 2010–2022 (ranging from 38779 to 224330 per disease cohort). We examined how often each diagnosed disease appeared on the certificate, its location and selection as the UCD, factors associated with its appearance (logistic regression), disease-specific mortality (multiple decrement life tables), and disease associations (Cause of Death Association Indicator, CDAI).

**Results:** Cancers appeared on the death certificate far more often than chronic diseases (around 75% versus 19–58%) and were usually recorded in Part 1 and selected as the UCD, whereas chronic diseases were rarely the UCD. The odds of a disease appearing depended on factors such as age at and time since diagnosis. When a diagnosed disease was recorded, the certificate traced a coherent path to death; when it was absent, ill-defined causes became more common. The CDAI highlighted specific association pathways between diseases.

**Conclusions:** MCoD data capture only part of the chronic disease burden present at death and should be interpreted cautiously as a proxy for end-of-life multimorbidity. They are, however, well suited to describing the pathways leading to death.

**Key messages:**

- Across COPD, dementia, diabetes, and cancer, multiple causes of death data capture only part of the chronic disease burden present at death and should be interpreted cautiously as a proxy for end-of-life multimorbidity.
- Cancers appear on most death certificates and are usually selected as the underlying cause, whereas chronic diseases appear far less often, making the underrepresentation of chronic conditions clear by contrast.
- When a diagnosed disease is recorded the certificate traces a coherent path to death, and the associations identified between conditions make these data well suited to describing the pathways leading to death rather than the overall burden of morbidity.

## Background

The process leading to death is often complex, involving numerous health risks and diseases that individuals face throughout their lifespan. However, mortality analysis often simplifies this process by focusing on the underlying cause of death (UCD), defined as the condition that initiated the sequence of events leading to death (1). While valuable for identifying the main health threats in a population, focusing solely on the UCD obscures the broader morbidity process leading to death. Moreover, because determining a single cause is difficult when multiple conditions are involved, misreporting or misclassification of the UCD has been a persistent issue in mortality studies (2,3).

In reality, most people aged 60+ are multimorbid and die with several diseases (4,5). Death certificates are designed to capture this complexity through a structured format with two parts: Part 1 records the causal sequence of events leading directly to death (immediate, intermediate, and underlying causes), while Part 2 records other significant conditions present at the time of death (6). The number of contributing causes recorded per death has been increasing over time and rises with age (7,8).

To offer a clearer picture of the morbid conditions leading to death, researchers have started investigating all the causes listed on the death certificate, referred to as Multiple Causes of Death (MCoD) analysis (9). MCoD data give deeper insight into competing causes and are especially valuable for highlighting conditions that, while not the primary cause, contribute to the mortality process, such as chronic diseases often relegated to secondary or contributory roles on the certificate (2,10,11).

MCoD data are therefore used as a proxy of the end-of-life health conditions (10,12–14). However, the likelihood that a disease is recorded on the death certificate is affected by factors beyond the individual’s health status, including place of death, educational attainment, and the certifying physician’s experience (14,15). This raises an important but largely underinvestigated question: how well do death certificate data actually represent the morbidity present at the end of life? A few record-linkage studies have begun to address this by comparing pre-death diagnoses with the causes recorded on the certificate, finding poor individual-level agreement that varies markedly by disease (16,17). Building on the MCoD literature and using high-quality Danish registry data, we address this by assessing whether recorded causes of death reflect end-of-life health conditions or rather the chain of events leading to death, drawing on individuals’ disease histories.

## Data and Methods

In our study, we used multiple high-quality Danish registers. The core data sources include the Danish Population Register, the Cause of Death Register, the Chronic Diseases and Severe Mental Disorders Register, and the Cancer Register. Using personal identifiers, we linked records from different registries for the same individual, enabling us to connect demographic information with detailed health diagnoses and cause-of-death data. We analysed data from 2010 onwards, when the Chronic Diseases registry reaches its highest quality (18): the registry applies a standardised look-back window to assess diagnostic criteria, and earlier years lack the full data horizon, potentially underestimating prevalence for some diseases.

The study population included all Danish residents aged 50 and older who had been diagnosed with chronic obstructive pulmonary disease (COPD), dementia (including Alzheimer’s disease), and diabetes type II. These diseases were selected because they can be tracked at the population level through the Chronic Diseases register, and because they represent highly prevalent conditions that are frequently underrepresented in conventional UCD analysis (19–21). We also analysed all cancers combined and five site-specific cancers — lung, breast, prostate, pancreatic, and colorectal — using the Danish Cancer Register. Cancers are often better represented in UCD analysis and can provide a benchmark against which to compare the other conditions. For each individual, we gathered information on their sex, date of birth, date and age of diagnosis, date and age at death, multiple causes of death, the role of the doctor recording the death, and highest educational attainment.

The analysis was divided into two parts: the first assessing whether MCoD data are representative of morbid conditions present at the end of life, and the second whether they capture the chain of events leading to death and disease-specific mortality patterns. All data management, statistical analyses, and visualisations were performed in R version 4.4.3 (22).

### Part 1: Representation of morbid conditions on death certificate

As a first exploration of the Danish MCoD data, we investigated, over the full study period 2010-2022, (1)* the number of times and percentage each diagnosed condition appears on the certificate (in any position), (2) its location on the certificate, among those for whom the condition is mentioned, (3) whether it is selected as the UCD, and (4) the average number of causes recorded for those with and without it reported. The UCD is determined using the Iris coding system, which incorporates the ACME (Automated Classification of Medical Entities) decision tables to select a single underlying cause from all conditions reported on the death certificate (23) (see Appendix for details on the Danish cause of death registration system).

To identify factors associated with the reporting of each diagnosed condition on the death certificate, we performed logistic regression models. The outcome variable captures whether the condition was reported on the death certificate (yes/no), while independent variables include: sex, age at diagnosis (50-64 years as reference, 65-79 years, 80+ years), time elapsed between diagnosis and death (0-1 year as reference, 1-2 years, 2-3 years, 3+ years), educational attainment (up to secondary as reference, tertiary, unknown or other), and doctor type recording the death (own doctor as reference, on-call doctor, hospital doctor, unknown or other).

### Part 2: Causes of death patterns and disease associations

Next, we examined the cause-of-death distribution of people with each diagnosed condition, comparing those who had their disease recorded on the certificate with those who did not. To do this, we computed multiple decrement life tables (MDLT) by sex (results for women are in the Appendix), 2-year periods (from 2010-11 to 2020-21), disease (COPD, dementia, diabetes, cancer), disease appearance on the death certificate (yes/no), and cause of death categories based on the Human Cause of Death Database (HCD) classification system (17 categories, see Appendix) (24). MDLT is a demographic technique that partitions mortality risk across competing causes of death. Confidence intervals were computed using bootstrap resampling with replacement and 1000 simulations (5000 for diabetes).

Finally, we assessed associations between each disease and the 17 HCD cause-of-death categories using the Cause of Death Association Indicator (CDAI) (25). The CDAI measures whether specific causes of death appear more or less frequently than expected — assuming independence across diseases — on death certificates when a particular disease is present, accounting for age structure through standardisation to the Danish population age-at-death distribution. Values above 100 indicate a positive association between the two conditions, and values below 100 a negative one.

For both analyses, we focused on COPD, dementia, diabetes, and cancer, but did not distinguish cancers by site due to insufficient deaths.

## Results

### Part 1: Representation of morbid conditions on death certificate

Table 1 displays the reporting probability of each disease on the death certificate among individuals diagnosed with that disease who died over the study period and the distribution of certificate locations where the disease appears. Reporting probability is highest for cancer (75%) compared to chronic diseases such as dementia (58%), COPD (37%), and diabetes (19%). Among the specific cancers examined, the highest reporting probabilities are for pancreatic and lung cancers (93% and 86% respectively) while breast cancer has the lowest (65%).

**Table 1.** Probability of each diagnosed disease being reported on the death certificate, and its location on the certificate, among diagnosed individuals aged 50+ who died, Denmark, 2010–2022.

| **Population with COPD** |  |  |
| --- | --- | --- |
| **Disease reported on death certificate** | **n** | **%** |
| No | 27930 | 62,9 |
| **Yes** | **16441** | **37,1** |
| **Placement on death certificate** | **n** | **%** |
| 1.a | 1052 | 6,4 |
| 1.b | 1986 | 12,1 |
| 1.c | 833 | 5,1 |
| 1.d | 3731 | 22,7 |
| 2.1 | 5672 | 34,5 |
| 2.2 | 2123 | 12,9 |
| 2.3 | 752 | 4,6 |
| 2.4 | 282 | 1,7 |
| **Population with dementia** |  |  |
| **Disease reported on death certificate** | **n** | **%** |
| No | 32032 | 42,4 |
| **Yes** | **43537** | **57,6** |
| **Placement on death certificate** | **n** | **%** |
| 1.a | 7550 | 17,3 |
| 1.b | 3720 | 8,5 |
| 1.c | 1287 | 3,0 |
| 1.d | 10939 | 25,1 |
| 2.1 | 14395 | 33,1 |
| 2.2 | 3777 | 8,7 |
| 2.3 | 1257 | 2,9 |
| 2.4 | 551 | 1,3 |

| Population with diabetes |  |  |
| --- | --- | --- |
| Disease reported on death certificate | n | % |
| No | 31430 | 81,0 |
| **Yes** | **7349** | **19,0** |

| Placement on death certificate | n | % |
| --- | --- | --- |
| 1.a | 25 | 0,3 |
| 1.b | 142 | 1,9 |
| 1.c | 142 | 1,9 |
| 1.d | 489 | 6,7 |
| 2.1 | 3433 | 46,7 |
| 2.2 | 2053 | 27,9 |
| 2.3 | 814 | 11,1 |
| 2.4 | 248 | 3,4 |

| Population with any cancer |  |  |
| --- | --- | --- |
| Disease reported on death certificate | n | % |
| No | 55545 | 24,8 |
| **Yes** | **168785** | **75,2** |

| Placement on death certificate | n | % |
| --- | --- | --- |
| 1.a | 53547 | 31,8 |
| 1.b | 21318 | 12,6 |
| 1.c | 10673 | 6,3 |
| 1.d | 64436 | 38,2 |
| 2.1 | 13662 | 8,1 |
| 2.2 | 3204 | 1,9 |
| 2.3 | 1282 | 0,8 |
| 2.4 | 519 | 0,3 |

| Population with lung cancer |  |  |
| --- | --- | --- |
| Disease reported on death certificate | n | % |
| No | 6322 | 14,2 |
| **Yes** | **38120** | **85,8** |

| **Placement on death certificate** | **n** | **%** |
| --- | --- | --- |
| 1.a | 10769 | 28,6 |
| 1.b | 4177 | 11,1 |
| 1.c | 1374 | 3,6 |
| 1.d | 18686 | 49,6 |
| 2.1 | 2087 | 5,5 |
| 2.2 | 372 | 1,0 |
| 2.3 | 129 | 0,3 |
| 2.4 | 63 | 0,2 |

| **Population with breast cancer** |  |  |
| --- | --- | --- |
| **Disease reported on death certificate** | **n** | **%** |
| No | 3761 | 34,8 |
| **Yes** | **7046** | **65,2** |

| **Placement on death certificate** | **n** | **%** |
| --- | --- | --- |
| 1.a | 1799 | 25,6 |
| 1.b | 585 | 8,3 |
| 1.c | 242 | 3,4 |
| 1.d | 2617 | 37,2 |
| 2.1 | 1229 | 17,5 |
| 2.2 | 341 | 4,9 |
| 2.3 | 149 | 2,1 |
| 2.4 | 66 | 0,9 |

| **Population with prostate cancer** |  |  |
| --- | --- | --- |
| **Disease reported on death certificate** | **n** | **%** |
| No | 4118 | 29,4 |
| **Yes** | **9891** | **70,6** |

| **Placement on death certificate** | **n** | **%** |
| --- | --- | --- |
| 1.a | 2393 | 24,2 |
| 1.b | 798 | 8,1 |
| 1.c | 284 | 2,9 |
| 1.d | 3942 | 39,9 |
| 2.1 | 1652 | 16,7 |
| 2.2 | 521 | 5,3 |
| 2.3 | 194 | 2,0 |
| 2.4 | 94 | 1,0 |

| **Population with colorectal cancer** |  |  |
| --- | --- | --- |
| **Disease reported on death certificate** | **n** | **%** |
| No | 6781 | 27,4 |
| **Yes** | **17998** | **72,6** |

| **Placement on death certificate** | **n** | **%** |
| --- | --- | --- |
| 1.a | 5026 | 28,2 |
| 1.b | 1642 | 9,2 |
| 1.c | 774 | 4,3 |
| 1.d | 8297 | 46,5 |
| 2.1 | 1468 | 8,2 |
| 2.2 | 411 | 2,3 |
| 2.3 | 147 | 0,8 |
| 2.4 | 76 | 0,4 |

| **Population with pancreatic cancer** |  |  |
| --- | --- | --- |
| **Disease reported on death certificate** | **n** | **%** |
| No | 792 | 7,0 |
| **Yes** | **10443** | **93,0** |

| **Placement on death certificate** | **n** | **%** |
| --- | --- | --- |
| 1.a | 3962 | 38,0 |
| 1.b | 933 | 8,9 |
| 1.c | 334 | 3,2 |
| 1.d | 4826 | 46,2 |
| 2.1 | 340 | 3,3 |
| 2.2 | 30 | 0,3 |
| 2.3 | 10 | 0,1 |
| 2.4 |  |  |

Regarding certificate location, diseases show distinct reporting patterns. Cancer is more often reported in Part 1 (in 75% or higher of cases where it appears). For COPD and dementia, we observe roughly equal distributions between Part 1 and Part 2. Finally, diabetes appears predominantly in Part 2 (approximately 80% of cases where it is recorded).

Table 2 displays whether each disease was selected as the UCD and how the average number of causes recorded differs by whether the diagnosed condition is reported. The ACME selection probability closely mirrors the Part 1/Part 2 distribution: for chronic diseases, 64% for dementia, 52% for COPD, and 19% for diabetes. In contrast, cancers show substantially higher ACME selection probability, with any cancer selected as UCD in 92% of cases where it appears, and pancreatic cancer reaching 97%. Breast and prostate cancers are lowest among cancers, at 80% and 82%. Furthermore, for chronic diseases, certificates listing the diagnosed condition record substantially more causes overall (e.g., 3.9 vs. 2.7 for COPD), whereas for cancers the difference is negligible (2.6 vs. 2.7 for any cancer).

**Table 2.** Probability of each diagnosed disease being selected as the underlying cause of death, and the mean number of causes recorded by whether the disease was reported, among diagnosed individuals aged 50+ who died, Denmark, 2010–2022.

| Population with COPD |  |  |
| --- | --- | --- |
| Disease reported in the ACME column | n | % |
| No | 7892 | 48,0 |
| **Yes** | **8549** | **52,0** |
| Average number of reported causes on death certificate if the disease is |  |  |
|  | n |  |
| Not reported | 2,7 |  |
| **Reported** | **3,9** |  |
| Population with dementia |  |  |
| Disease reported in the ACME column | n | % |
| No | 15803 | 36,3 |
| **Yes** | **27734** | **63,7** |
| Average number of reported causes on death certificate if the disease is |  |  |
|  | n |  |
| No | 2,5 |  |
| **Yes** | **3,1** |  |
| Population with diabetes |  |  |
| Disease reported in the ACME column | n | % |
| No | 5969 | 81,2 |
| **Yes** | **1380** | **18,8** |

| Average number of reported causes on death certificate if the disease is | n |
| --- | --- |
| No | 2,8 |
| **Yes** | **4,5** |

| Population with any cancer |  |  |
| --- | --- | --- |
| Disease reported in the ACME column | n | % |
| No | 12905 | 7,6 |
| **Yes** | **155880** | **92,4** |

| Average number of reported causes on death certificate if the disease is | n |
| --- | --- |
| Not reported | 2,7 |
| **Reported** | **2,6** |

| Population with lung cancer |  |  |
| --- | --- | --- |
| Disease reported in the ACME column | n | % |
| No | 2220 | 5,8 |
| **Yes** | **35900** | **94,2** |

| Average number of reported causes on death certificate if the disease is | n |
| --- | --- |
| No | 2,5 |
| **Yes** | **2,5** |

| Population with breast cancer |  |  |
| --- | --- | --- |
| Disease reported in the ACME column | n | % |
| No | 1409 | 20,0 |
| **Yes** | **5637** | **80,0** |

| Average number of reported causes on death certificate if the disease is | n |
| --- | --- |
| No | 2,6 |
| Yes | 2,8 |

| Population with prostate cancer |  |  |
| --- | --- | --- |
| Disease reported in the ACME column | n | % |
| No | 1814 | 18,3 |
| Yes | 8077 | 81,7 |

| Average number of reported causes on death certificate if the disease is | n |
| --- | --- |
| Not reported | 2,6 |
| Reported | 2,9 |

| Population with colorectal cancer |  |  |
| --- | --- | --- |
| Disease reported in the ACME column | n | % |
| No | 1559 | 8,7 |
| Yes | 16439 | 91,3 |

| Average number of reported causes on death certificate if the disease is | n |
| --- | --- |
| No | 2,7 |
| Yes | 2,6 |

| Population with pancreatic cancer |  |  |
| --- | --- | --- |
| Disease reported in the ACME column | n | % |
| No | 278 | 2,7 |
| Yes | 10165 | 97,3 |

| Average number of reported causes on death certificate if the disease is | n |
| --- | --- |
| No | 2,1 |
| Yes | 2,3 |

Figure 1 presents the odds ratios from logistic regression models examining factors associated with disease appearance on death certificates among individuals diagnosed with each condition. Table A1 in the Appendix presents the full model results. Sociodemographic characteristics (i.e., sex and educational attainment) appear to play only a minor role in whether diseases are reported, while three factors show stronger associations: time elapsed between diagnosis and death (positive for chronic diseases, negative for cancers); age at diagnosis (older age linked to lower reporting for cancers and COPD, higher for dementia and diabetes); and doctor type (hospital doctors showing notably higher odds for COPD than the patient’s own doctor).

**Figure 1.**
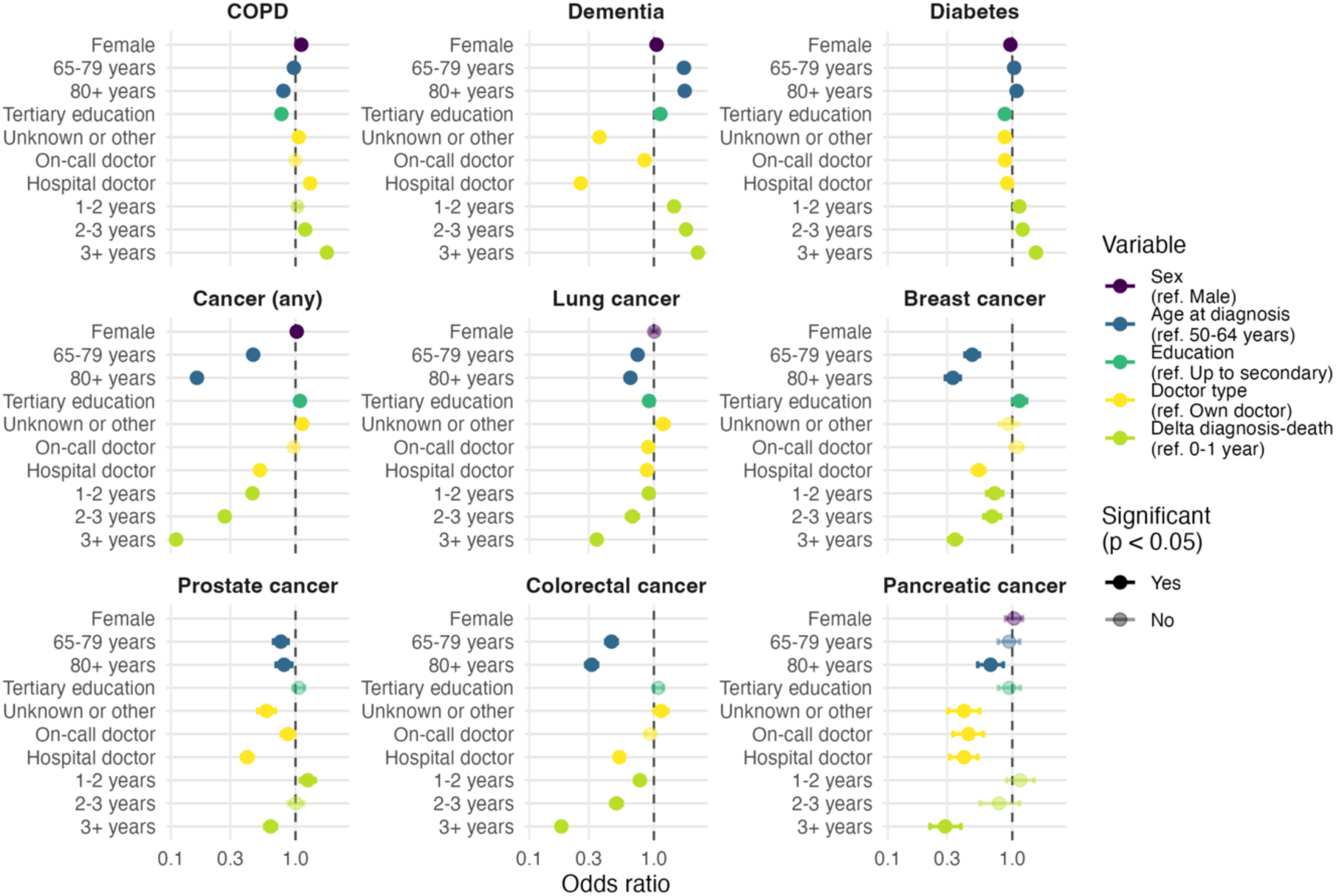
Adjusted odds ratios for factors associated with each diagnosed disease (COPD, dementia, diabetes, and cancer) being reported on the death certificate, Denmark, 2010-2022.

### Part 2: Causes of death patterns and disease associations

Figures 2-5 present the proportion of deaths by the four leading causes over time for men in each disease subpopulation, stratified by whether the diagnosed disease appears on the death certificate. As expected, individuals diagnosed with a specific disease are more likely to die from it than the general population; for example, individuals with COPD more frequently die from COPD or other respiratory diseases.

**Figure 2.**
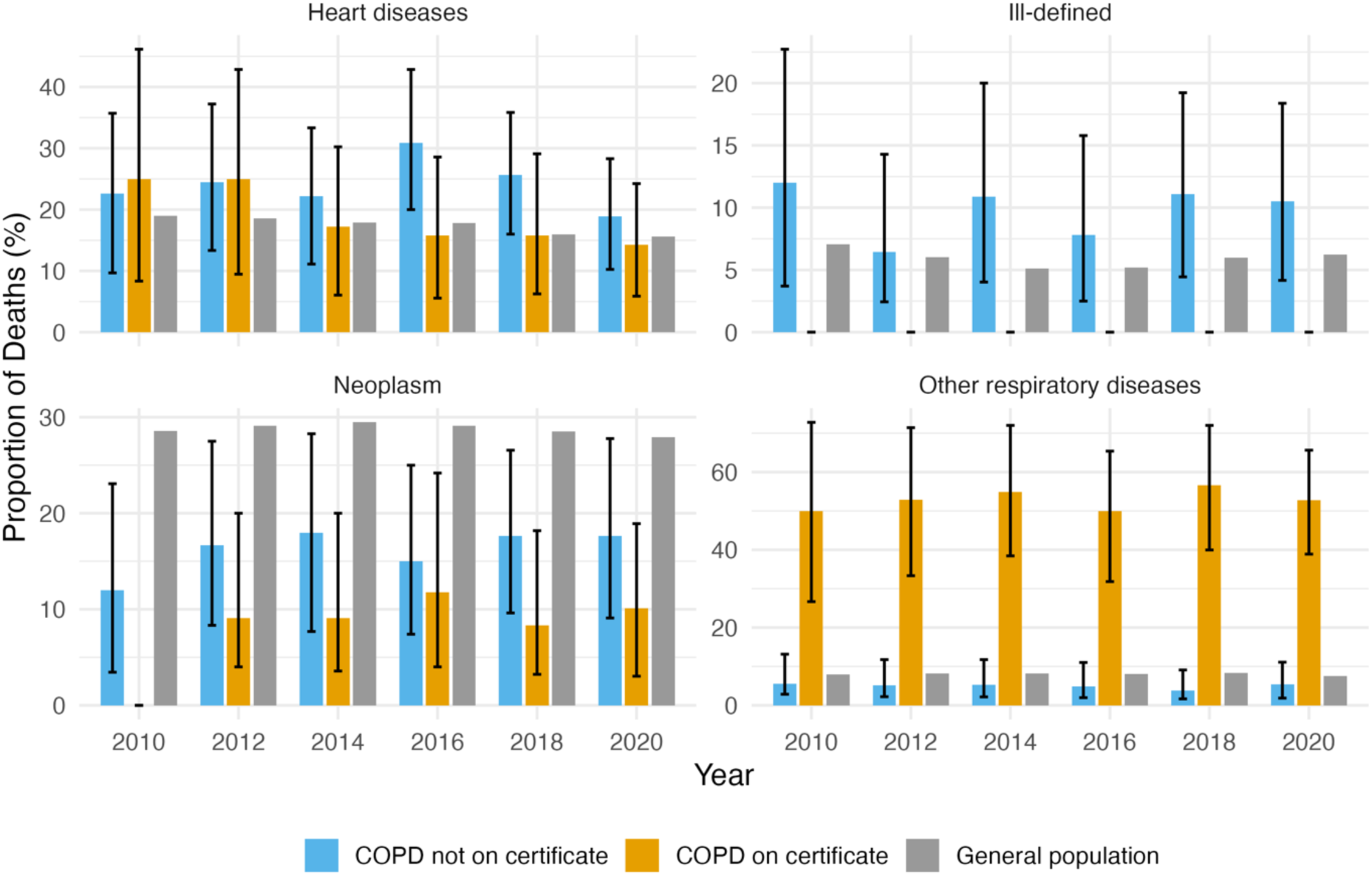
Proportion of deaths by the four leading causes of death over time among men with COPD, by whether COPD was reported on the death certificate, with the general male population for comparison, Denmark, 2010-2021.

**Figure 3.**
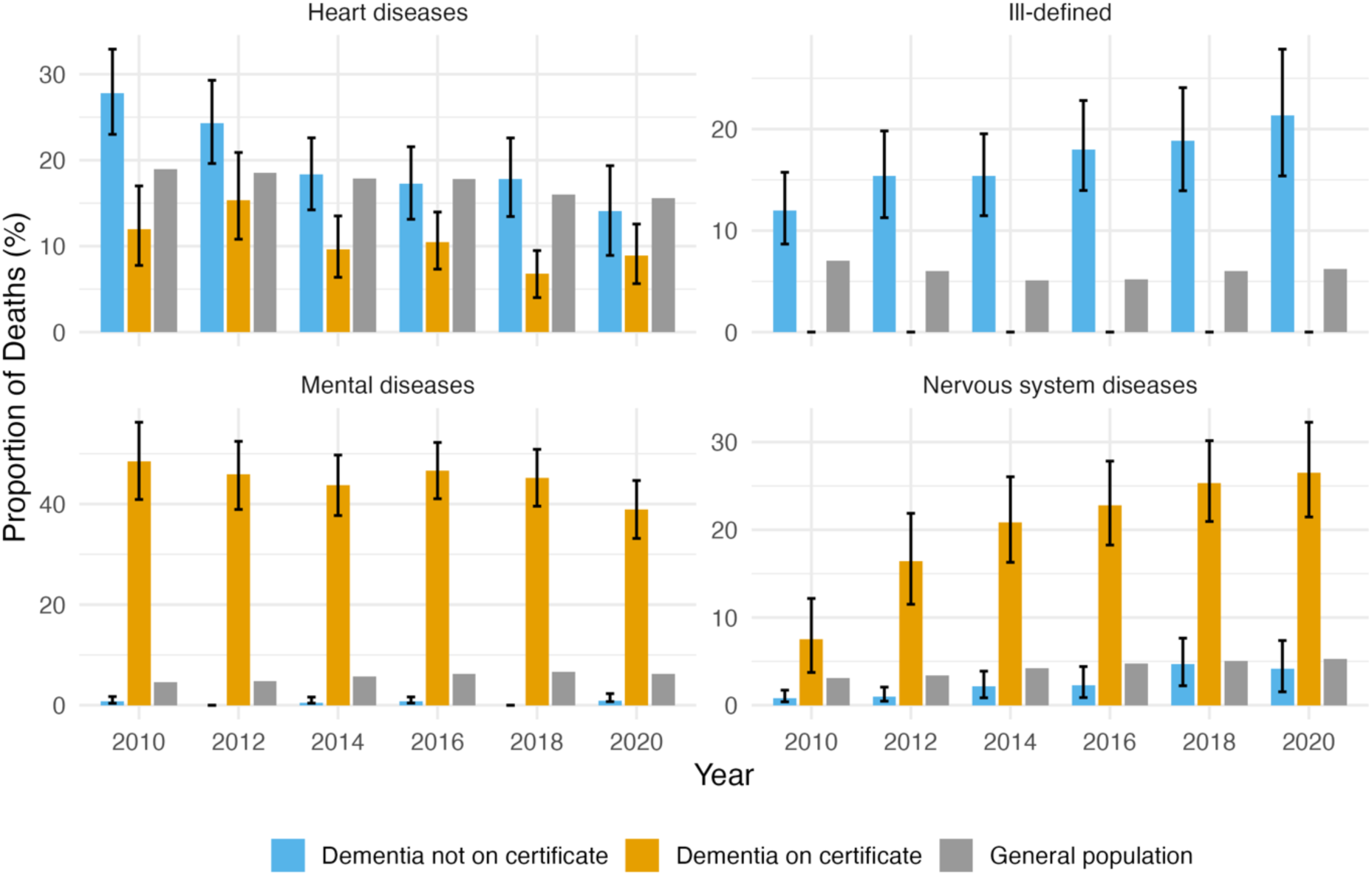
Proportion of deaths by the four leading causes of death over time among men with dementia, by whether dementia was reported on the death certificate, with the general male population for comparison, Denmark, 2010-2021.

**Figure 4.**
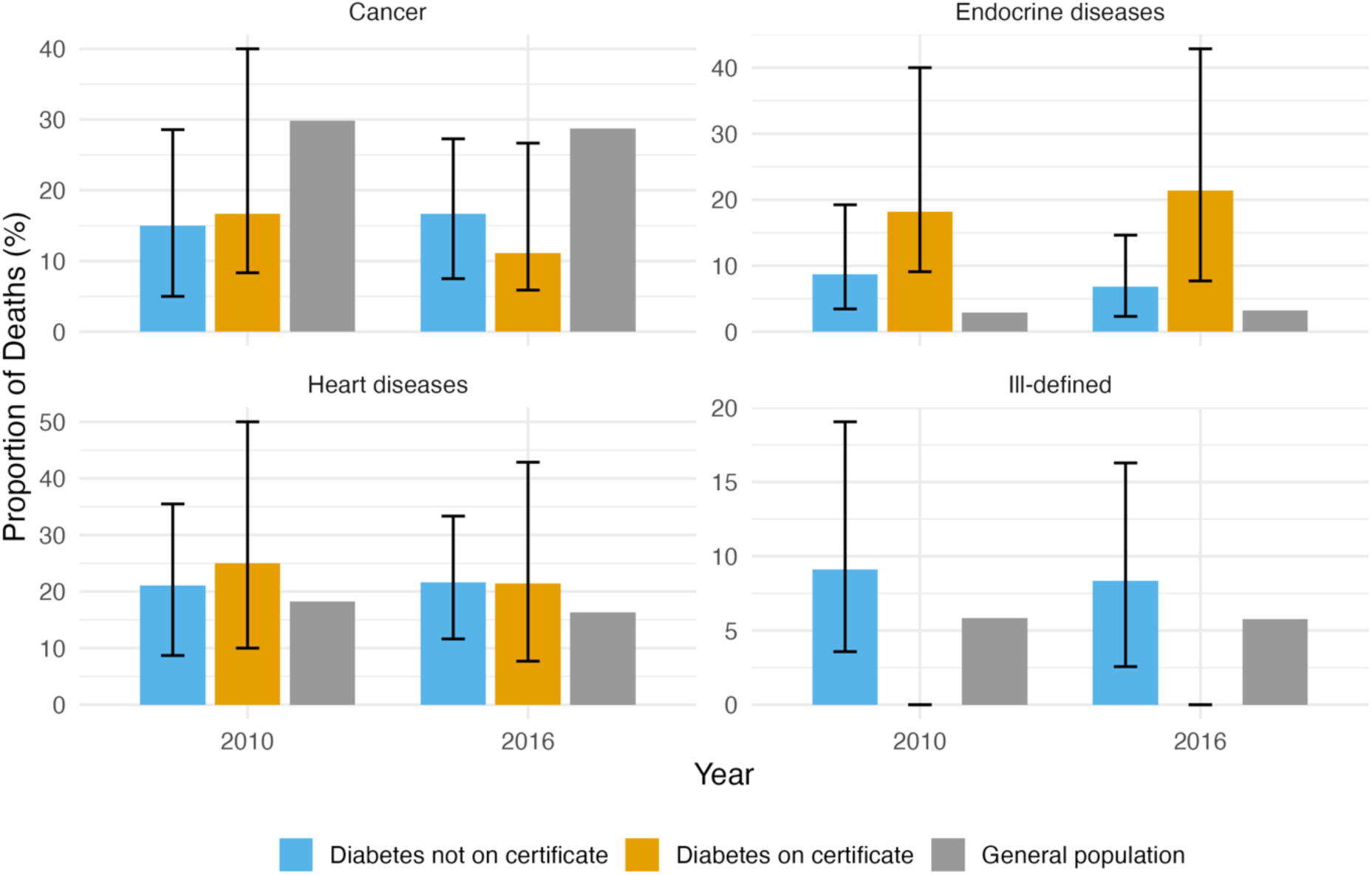
Proportion of deaths by the four leading causes of death over time among men with diabetes, by whether diabetes was reported on the death certificate, with the general male population for comparison, Denmark, 2010-2021.

**Figure 5.**
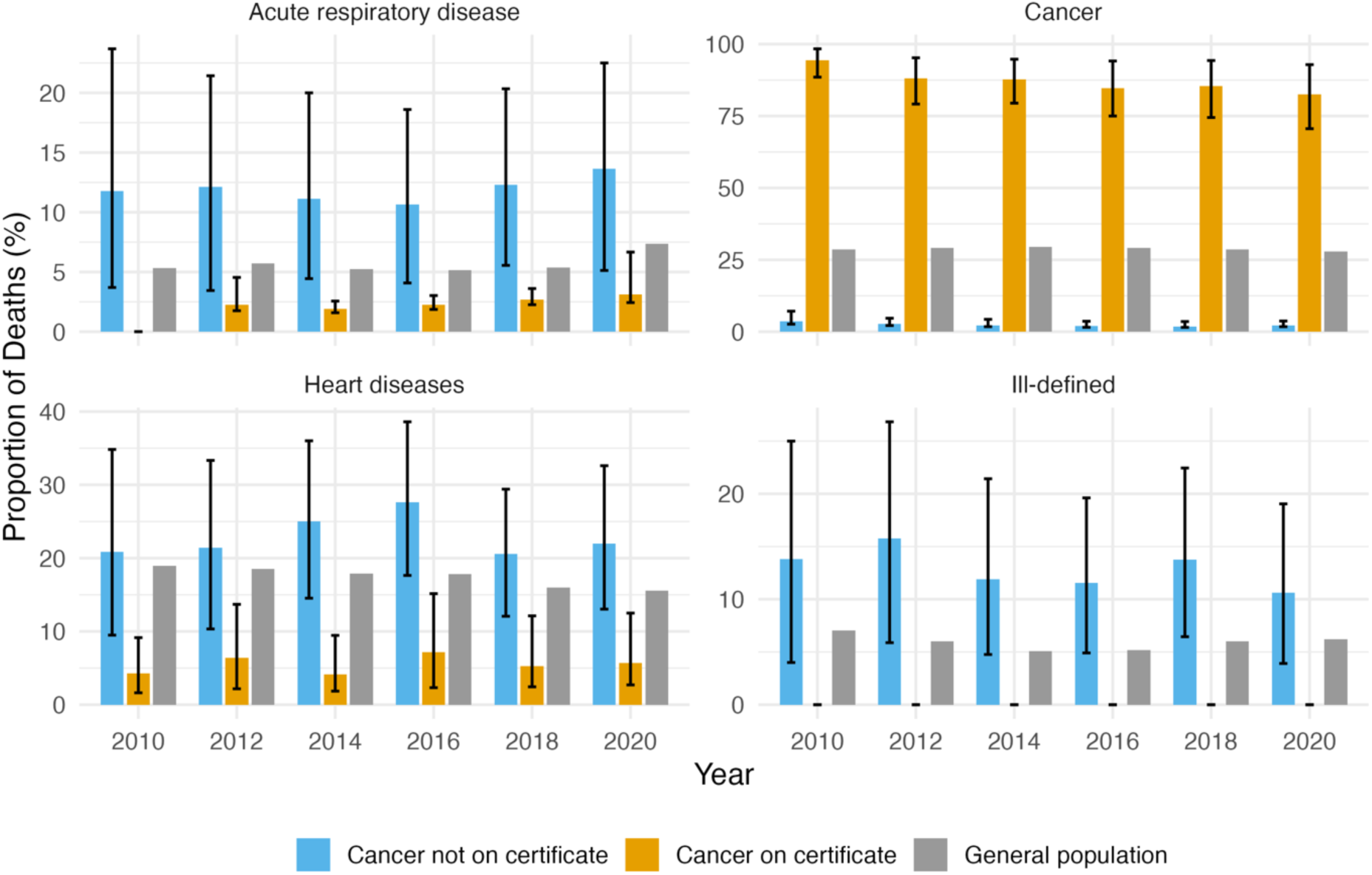
Proportion of deaths by the four leading causes of death over time among men with cancer, by whether cancer was reported on the death certificate, with the general male population for comparison, Denmark, 2010-2021.

However, the UCD varies depending on whether the diagnosed disease is reported. For most diseases examined, when it is not reported, individuals are more likely to have heart diseases or ill-defined causes recorded as their UCD — both compared to the general population and to those for whom it is reported.

For those without the diagnosed disease reported, the probability of having cancer as the UCD is higher than among those whose disease is reported, but still lower than in the general population.

Figure 6 presents the CDAI (with 95% confidence intervals) for the association between each diagnosed disease, when reported as a contributory cause, and the underlying cause of death. The y-axis displays causes of death ordered by mean CDAI values across all diseases. Points are coloured according to association strength, with darker red indicating stronger associations (more standard deviations above the reference value of 100). Grey points (at CDAI = 100) mark disease-specific cause combinations excluded from the analysis, as these correspond to the diagnosed conditions themselves or their direct complications.

**Figure 6.**
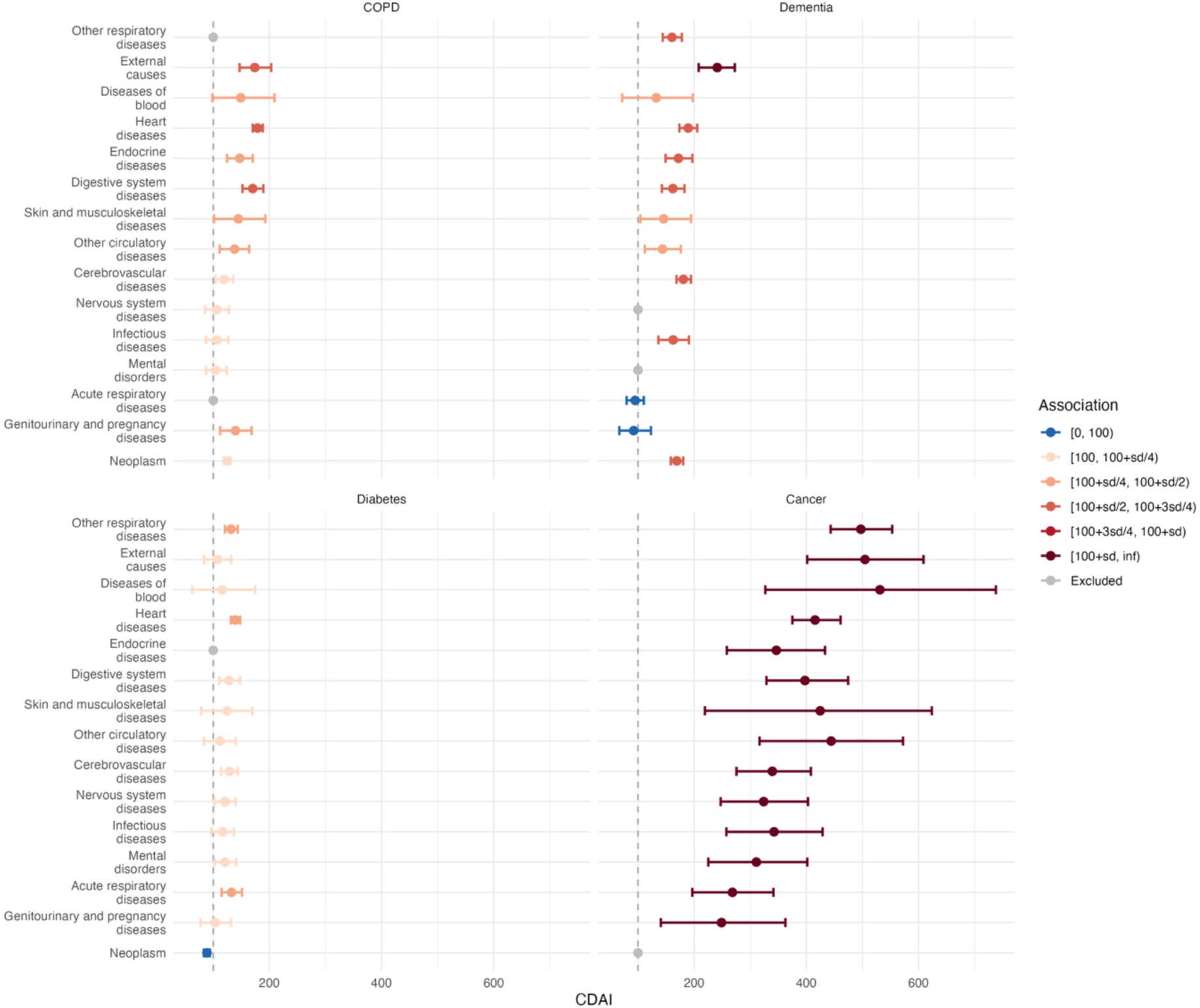
Cause of Death Association Indicator (CDAI) between each diagnosed disease, when reported as a contributory cause, and the underlying causes of death, Denmark, 2010-2022 (points show CDAI values with 95% confidence intervals).

The plot reveals distinct association patterns across diseases. Cancer diagnoses show the strongest associations with all other causes of death (CDAI > 200), though confidence intervals are very large due to the small number of cases where cancer is a contributing cause. In contrast, COPD shows relatively weak associations with most causes, except heart diseases, external causes, and digestive system diseases, where CDAI values approach 150-200 with relatively small confidence intervals. Dementia exhibits strong associations primarily with external causes (CDAI > 200) but also, to a lesser extent, with heart, cerebrovascular, other respiratory, endocrine, infectious and digestive diseases, as well as neoplasms. Diabetes shows the weakest associations, with only moderate ones for heart, acute and other respiratory diseases.

## Discussion

Using comprehensive Danish registry data linking disease diagnoses to death certificates, this study examined how frequently diagnosed conditions translate into recorded causes of death and how cause-of-death profiles differ across morbid populations. Overall, MCoD data poorly capture end-of-life morbidity: chronic conditions such as COPD and diabetes are frequently absent from the certificate, whereas cancer is markedly more likely to be recorded as a cause of death.

### MCoD data as proxy of multimorbidity at the end of life

Our findings quantify the gap between chronic disease burden in morbidity data and that recorded on death certificates, consistent with previous validation studies (16,26–28). Only a minority of individuals diagnosed with COPD, dementia, or diabetes had these conditions recorded anywhere on their death certificates, contrasting sharply with cancers. Whilst not all diagnosed conditions should appear on the certificate, only those contributing to death, MCoD data are nonetheless frequently used as proxies for end-of-life multimorbidity (10,12–14), a practice our results show produces an incomplete picture of chronic disease burden at death.

We examined how demographic and clinical characteristics relate to disease reporting, though these patterns likely also reflect differences in cause-of-death profiles rather than reporting biases alone. Time elapsed between diagnosis and death showed the strongest associations: for chronic diseases, longer time since diagnosis increased reporting, whereas for cancers shorter times did. Neoplasms often advance rapidly and are recorded as the UCD shortly after diagnosis, typically with few competing conditions; with longer survival, a cancer may instead behave like a chronic disease and is less likely to appear on the certificate. Diabetes and dementia, by contrast, progress gradually, so their contribution to mortality accumulates over time before being recorded.

Age at diagnosis and educational attainment — the latter with odds ratios very close to 1 — also showed significant associations with disease reporting. Educational attainment is protective against both diabetes and COPD incidence (29,30), while the highest prevalence and incidence of chronic diseases, including dementia, are observed at older ages (31).

The certifying physician also mattered: the patient’s own doctor showed higher odds of reporting chronic diseases than other physicians, with the exception of COPD, which hospital doctors reported more often. This likely reflects three mechanisms: COPD frequently requires hospital care for acute exacerbations; access to records differs, with own doctors knowing patients’ chronic conditions while hospital doctors document the acute respiratory events leading to death; and disease severity, as those dying in hospital present more advanced cases where COPD more directly precipitates death.

### Causes of death patterns and disease associations

The multiple decrement life tables revealed markedly different cause-of-death profiles depending on whether the diagnosed disease appeared on the certificate. When recorded, deaths were predominantly attributed to causes directly related to the condition: respiratory diseases for COPD, mental and nervous system disorders for dementia, endocrine and heart diseases for diabetes, and neoplasms for cancer. When absent, heart diseases and ill-defined causes were consistently more prevalent across all subpopulations. Most of these ill-defined deaths are coded as "unknown causes of death," which occur more frequently in Denmark than in many other countries (32), suggesting that when a diagnosed disease is not recorded, practitioners were often unable to identify the pathway leading to death and thus the underlying cause.

The elevated probability of heart disease as the UCD when the diagnosed disease is unreported may reflect both the well-known association between many chronic conditions and heart disease, and a competing-risk mechanism: when an individual has both the diagnosed condition and heart disease, the latter is more likely to be selected as the UCD (17).

The CDAI analysis points to meaningful mechanisms behind these associations. The link between COPD and heart disease is well established: COPD patients have a substantially increased risk of cardiovascular disease (33), driven by shared mechanisms including systemic inflammation, oxidative stress, and hypoxemia that promote atherosclerosis and cardiac dysfunction independently of shared risk factors like smoking (34). The association with digestive system diseases may reflect shared inflammatory pathways or medication effects, while the co-occurrence with external causes likely captures deaths where COPD contributed to vulnerability without being the primary cause.

For dementia, the association with external causes, particularly falls, reflects the well-documented elevated risk of unintentional injuries in dementia patients (35). The associations with heart and cerebrovascular diseases are consistent with the bidirectional relationship between vascular disease and dementia, whereby stroke and cardiovascular disease are both risk factors for and consequences of cognitive decline; vascular dementia shows particularly strong associations with cardiovascular mortality compared to other subtypes (36).

For diabetes, the association with heart disease reflects the well-established link between the two, with cardiovascular disease highly prevalent in type 2 diabetes and accounting for a substantial proportion of diabetes-related deaths (37). The associations with acute and other respiratory diseases are consistent with evidence that diabetes increases the risk of pneumonia, COPD, and respiratory failure (38).

Unlike the chronic conditions, whose associations were weaker, cancer diagnoses showed associations with all causes of death. Because cancer is usually the UCD rather than a contributing condition, these results rest on few cause-of-death combinations; nevertheless, the associations that emerge are meaningful. The co-occurrence of cancer with blood diseases, other respiratory diseases, and external causes likely reflects the systemic effects of cancer and its treatments, which can compromise haematopoietic function, respiratory capacity, and overall vulnerability to injury and infection. Overall, our results confirm the value of MCoD data for understanding pathways to death (39).

## Limitations

This study has several limitations. The relatively small sample sizes for specific disease-cause combinations precluded more detailed stratified analyses by period or age. Our analysis focused on three chronic conditions and selected cancers, which may not be representative of reporting patterns for other diseases. The restriction to deaths from 2010 onwards, whilst ensuring optimal register quality, limited our ability to assess longer-term trends in death certificate completion. Additionally, whilst we interpreted observed associations as reflecting genuine differences in cause-of-death profiles, we cannot entirely exclude the possibility that systematic reporting biases contribute to these patterns. A further limitation is selection: because the chronic conditions are ascertained less completely than cancer, the diagnosed cohorts may over-represent more severe cases, affecting both appearance proportions and the cancer-chronic contrast. Finally, our findings are specific to Denmark, where death certificate completion practices and access to comprehensive medical records may differ from other settings.

## Conclusion

Our findings demonstrate that MCoD data cannot serve as reliable proxies for multimorbidity at the end of life, capturing only part of the chronic disease burden present at death.

However, how and when conditions are reported is not random, as it reflects meaningful associations between diseases within the mortality process. While the quality of UCD data is often questioned, the MCoD approach can enhance our understanding of the pathways leading to death by accounting for multiple, often interconnected, conditions. Researchers and policymakers should recognize this distinction when interpreting MCoD data: whilst they do not capture overall population health status, these data are meant to document the chain of events leading to death.

## Declarations

### Ethics approval

Not applicable.

## Acknowledgements

The authors wish to thank Aleksandrs Aleksandrovs for his work on data management.

## Author contributions

C.S. and M.-P.B.-B. conceptualised the study. C.S. curated the data. C.S. and E.U. performed the analysis. C.S. wrote the original draft. All authors interpreted the results, contributed to the writing, and approved the final version to be published.

## Supplementary data

Supplementary data (Appendix) are available at IJE online.

## Data availability

The authors do not have permission to share data.

## Use of artificial intelligence (AI) tools

The authors used Claude (Anthropic) to assist with language editing and with preparing code to analyse the data and visualise the results. The authors reviewed and verified all content and take full responsibility for the integrity and accuracy of the work.

## Appendix

## Appendix 1: Death certification in Denmark

When a person in Denmark dies, a doctor conducts a post-mortem examination. The doctor then fills out a death certificate, which aligns with a standard WHO (2016) death certification form:

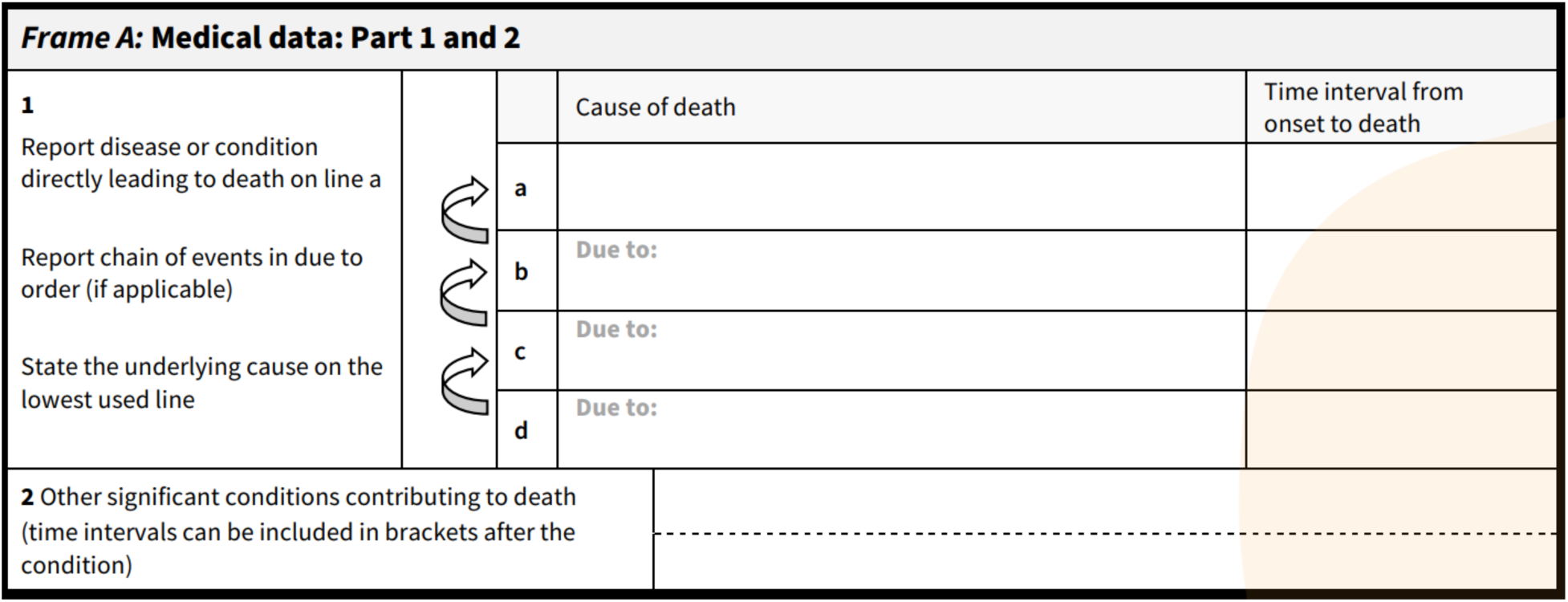

Source: WHO (2016)

In Part 1, the physician documents the sequence of morbid events that led to death. The earlier a condition began, the lower it should be recorded. The initiating conditions, referred to as the underlying cause of death, should be recorded on the lowest line. The immediate condition should occupy the highest line, while any intermediate conditions should be recorded in between.

In Part 2, the physician records contributory causes that are not directly related to the underlying cause. These should include other significant conditions that contributed to the patient’s death.

Filled form is then submitted electronically to the National Board of Health (NCRR, 2025). The data is then electronically transferred to the National Death Registry and processed centrally on national level (Helweg-Larsen, 2011).

In 2002, the automation of COD classification was introduced, and Denmark began using ACME decision tables. Later, in 2007, another innovation was implemented: full responsibility for COD certification was given to the physician performing the post-mortem examination, replacing the death coders who had previously conducted the central validation of the data received from the physicians. From that point onward, death certificates have been submitted electronically to the National Board of Health. Since 2012, Danish authorities have been using the Iris system as a coding aid (Helweg-Larsen, 2011; Iris, 2025).

## Appendix 2: Classification of causes of death into 17 categories, based on the short list of the Human Cause-of-Death Database (HCD), with corresponding ICD-10 codes.

| No. | Cause-of-death category | ICD-10 codes |
| --- | --- | --- |
| 1 | Infectious diseases | A00–B99 |
| 2 | Neoplasm | C00–D48 |
| 3 | Diseases of blood | D50–D89 |
| 4 | Endocrine diseases | E00–E88 |
| 5 | Mental disorders | F01–F99 |
| 6 | Nervous system diseases | G00–G44, G47–H93 |
| 7 | Heart diseases | I00–I51 |
| 8 | Cerebrovascular diseases | G45, I60–I69 |
| 9 | Other circulatory diseases | I70–I99, K64 |
| 10 | Acute respiratory diseases | J00–J22, U04, U07–U10 |
| 11 | Other respiratory diseases | J30–J98 |
| 12 | Digestive system diseases | K00–K63, K65–K92 |
| 13 | Skin and musculoskeletal diseases | L00–M99 |
| 14 | Genitourinary and pregnancy diseases | N00–O99 |
| 15 | Perinatal and congenital | P00–Q99, R95 |
| 16 | External causes | V01–Y89 |
| 17 | Ill-defined causes | R00–R94, R96–R99 |

## Appendix 3: Multiple Decrement Life Table results for women

**Figure A1.**
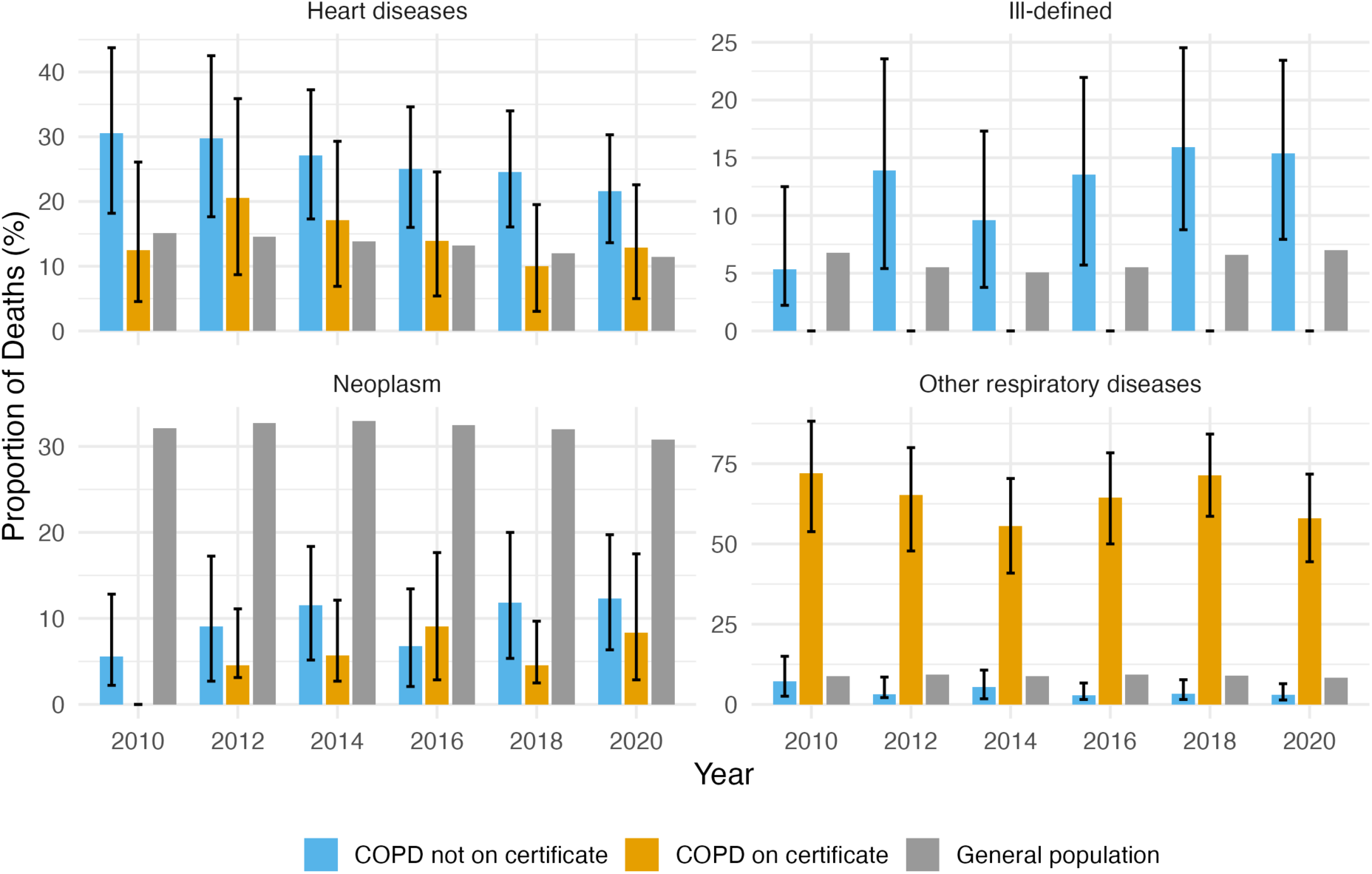
Proportion of deaths by the four leading causes of death over time among women with COPD, by whether COPD was reported on the death certificate, with the general female population for comparison, Denmark, 2010-2021.

**Figure A2.**
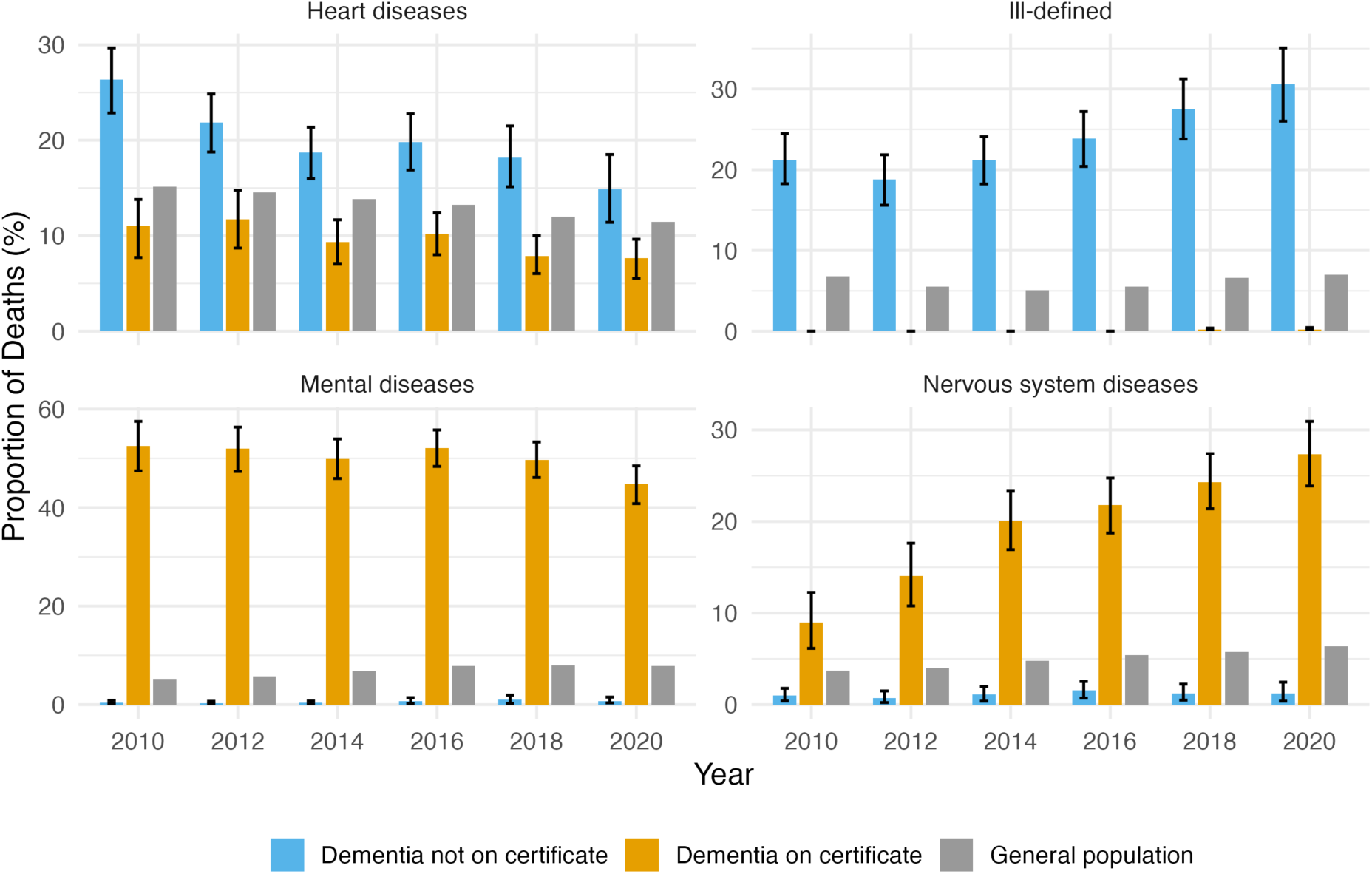
Proportion of deaths by the four leading causes of death over time among women with dementia, by whether dementia was reported on the death certificate, with the general female population for comparison, Denmark, 2010-2021.

**Figure A3.**
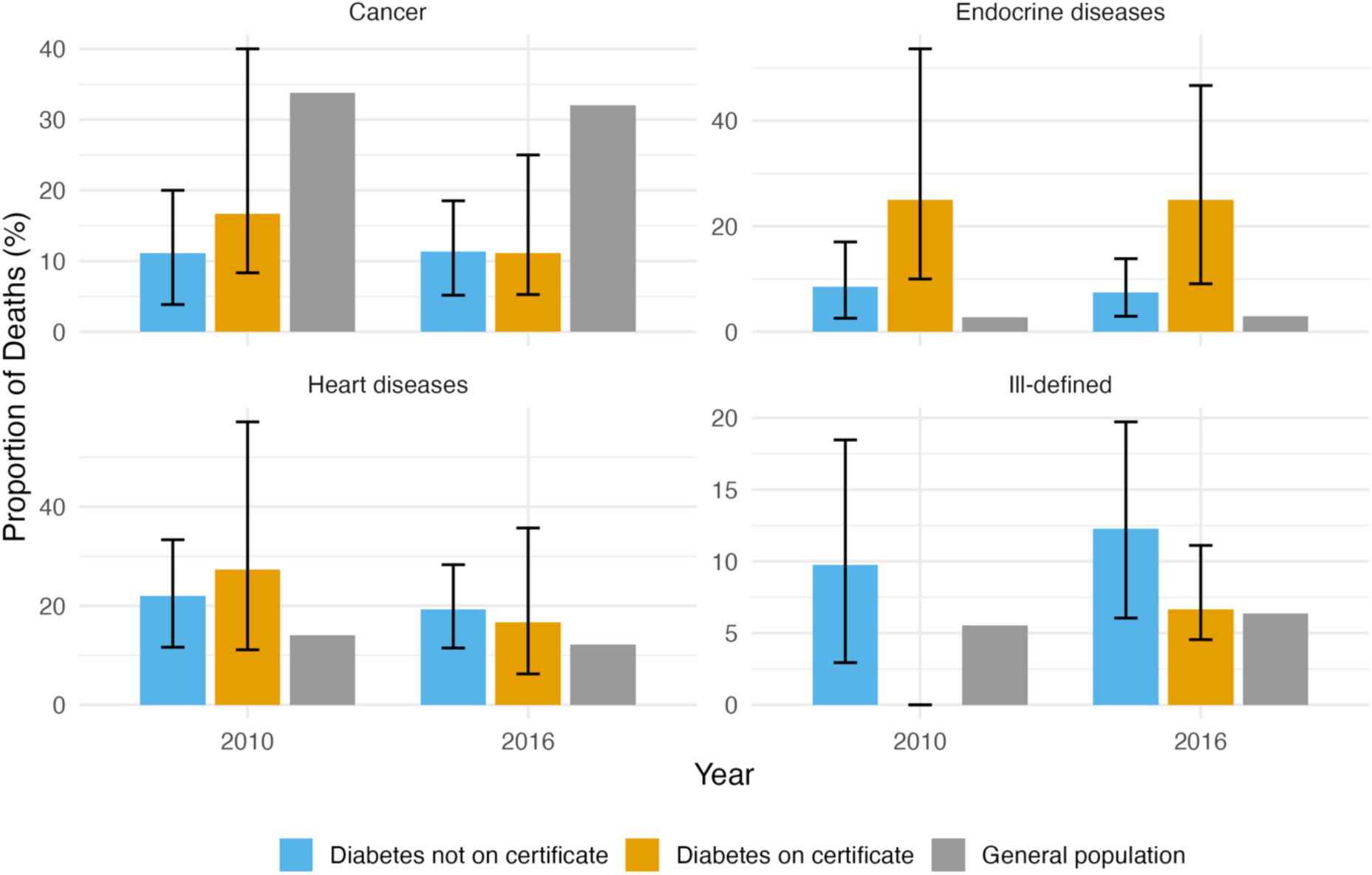
Proportion of deaths by the four leading causes of death over time among women with diabetes, by whether diabetes was reported on the death certificate, with the general female population for comparison, Denmark, 2010-2021.

**Figure A4.**
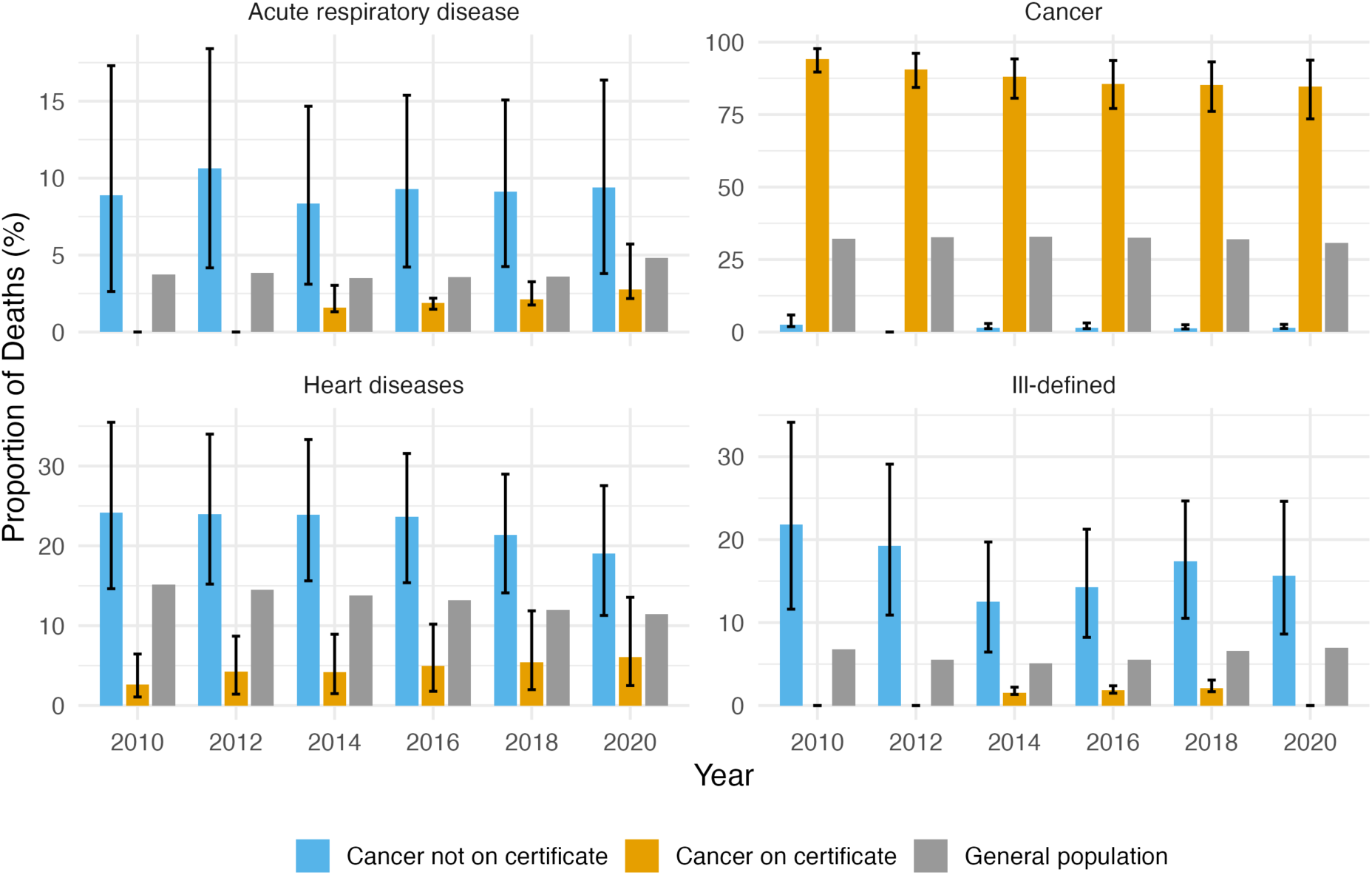
Proportion of deaths by the four leading causes of death over time among women with cancer, by whether cancer was reported on the death certificate, with the general female population for comparison, Denmark, 2010-2021.

## Appendix 4: Cause of death associations indicator

The Cause of death association indicator (CDAI) is calculated using the formula (Bishop et al., 2023) :

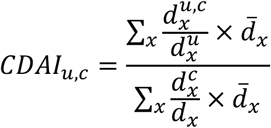

Where *d_x_^u,c^* is number of deaths at age x with UCD u and contributory CC. Where *d*^!^ is number of deaths at age x and cause u as UCD.

Where *d_x_^u^* is number of deaths at age x with cause c as contributory (regardless of underlying).

Where *d_x__c_* is number of deaths at age x.

Where *d̅_x_* is the standard population.

When the CDAI is not significantly different from one (or 100), it can be interpreted that the probability of dying with both diseases is the same as the probability of dying with either one. Dessesquelles et al. (2010) state that “*CDAI is the ratio of the standardized observed prevalence measure to the standardized expected prevalence measure assuming independence between diseases*” (Dessesquelles et al., 2010).

